# Semantic Memory in Healthy Apolipoprotein ε4 Carriers: A Systematic Review

**DOI:** 10.1101/2025.03.07.25323557

**Authors:** Riccardo Sacripante, Tabitha James, Michael Hornberger, Joshua Blake, Louis Renoult

## Abstract

The Apolipoprotein epsilon 4 (*APOE* ε4) genetic variant is notoriously linked to enhanced risk of developing Alzheimer’s Disease (AD). Several studies have examined how this allele could influence cognitive functioning in healthy adults, and whether ε4 carriers show a subtle cognitive decline that would indicate preclinical AD pathology. Research has predominantly focused on episodic memory, where ε4 carriers are usually impaired, while semantic memory functioning has received less attention. To evaluate current evidence on the influence of *APOE* ε4 on semantic memory, we systematically reviewed the research literature assessing semantic memory in non-clinical adult populations according to the PRISMA guidelines. We reviewed 17 studies that revealed high heterogeneity in how semantic memory is conceptualised and assessed. When tested via standard neuropsychological tests (i.e., category fluency, naming, language comprehension, and general knowledge), ε4 carriers did not significantly differ from non-carriers. Instead, ε4 carriers showed lower performance than non-carriers when assessed via more complex semantic memory tasks (i.e., longer category fluency tasks, autobiographical memory tasks, measures of semantic clustering). The impact of *APOE* ε4 on semantic memory thus appears to be restricted to these more complex tasks, which could constitute a better match to episodic memory tasks for which *APOE* effects are typically observed, though a mediating role of executive functions should also be considered. Future research investigating autobiographical memory retrieval in ε4 carriers could provide a more sensitive and ecologically valid assessment of semantic memory and would help disentangle personal and general forms of semantic memory.

## 1. Introduction

Alzheimer’s Disease (AD) is the most common form of neurodegenerative disease and dementia in the world, and it has become one of the most expensive and burdening conditions of this century (Scheltens et al., 2021). Early and accurate detection of AD is important for the screening, diagnosis and subsequent management and care of people affected by this neurodegenerative condition (Porsteinsson et al., 2021). However, detecting early deficits in preclinical AD is problematic and clinically difficult, given the vast heterogeneity of normal ageing and AD expression (Emrani et al., 2020). Early cognitive deficits often involve spatial navigation and episodic memory (Coughlan et al., 2018) and once a person receives a diagnosis, cognitive impairments are often fairly pronounced. Late-onset AD can therefore elude clinical detection for years and even decades, and this inevitably has a life-changing impact on the quality of life of people receiving such diagnosis and their families and carers (Rasmussen & Langerman, 2019). With the recent approval and imminent rollout of the first disease-modifying pharmacological treatments for AD (e.g., Donanembad, or Lecanemab; Mintun et al., 2021, see also Laurell et al., 2024), early detection of subtle cognitive markers of AD has become even more important.

Advances in neuroimaging measures like Positron Emission Tomography (PET), fluorodeoxyglucose PET (FDG-PET), or functional Magnetic Resonance Imaging (fMRI) (for a review see Ewers et al., 2011) in conjunction with AD biomarkers (e.g., beta-amyloid and tau proteins) have dramatically improved the precision of the AD diagnostic criteria (see McKhann et al., 2011 for AD). Indeed, changes in brain biochemistry involving biomarkers are now thought to occur approximately 20 years before the onset of classic AD symptoms (Alzheimer’s Association, 2019). In this regard, a promising ground of research derives from cognitive and genetic markers in preclinical AD which, along with brain biomarkers and sensitive cognitive assessment, could predict the development of the disease and inform future pharmacological and cognitive interventions (for a review see Jackson et al., 2024).

### 1.1 Apolipoprotein Epsilon 4 (*APOE* ε4)

*APOE*, or apolipoprotein E, is a protein that transports cholesterol and other fatty substances within brain cells and supplies the central nervous system with essential lipids. *APOE* corresponds to different versions of a DNA sequence on chromosome 19, known as an allele, with three major variants or isoforms (ε2, ε3, and ε4), for which every individual inherits one from each parent. Variants in allele genotypes can be homozygous (ε2ε2, ε3ε3, ε4ε4) or heterozygous (ε2ε3, ε2ε4, ε3ε4) and each isoform of the *APOE* protein corresponds to distinct structural properties which impact brain function.

It has been demonstrated that people carrying the ε4 variant of the *APOE* gene are at increased risk of developing sporadic late-onset Alzheimer’s Disease (Corder et al., 1993; Farrer et al., 1997) with an earlier age of onset (Fortea et al., 2024), while those carrying the ε2 allele are at a decreased risk (Reiman et al., 2020, for a review see Suri et al., 2013). Notably, ε4 homozygotes carriers (ε4ε4) present with greater risk compared to ε4 heterozygotes carriers (ε3ε4 or ε2ε4), meaning that genetic risk to AD could be dose-dependent (Blacker et al., 1997; Davidson et al., 2006). Despite the presence of the *APOE* ε4 genotype being restricted to only 20 to 25% of the general population in different global regions, the allele is highly present in cases of late-onset AD (i.e., almost half of all cases, see Caselli & Reiman, 2012). A recent study examining clinical, pathological, and biomarker changes in homozygotic *APOE* ε4 carriers (Fortea et al., 2024) concluded that this allele mutation represents a direct cause of late-onset AD and not just a risk factor, as almost all these participants presented with AD brain pathology already from middle age (see also Xu et al., 2024). It should, however, be noted that having high amyloid burden does not necessarily translate to AD (for a meta-analysis see Jansen et al., 2015).

A plethora of research studies focused their attention on how this allele could influence cognition and cognitive decline in non-demented healthy adults (see O’Donoghue et al., 2018; Small et al., 2004; Wisdom et al., 2011). Meta-analyses on the effect of *APOE* on cognition (Small et al., 2004; Wisdom et al., 2011) observed that *APOE* ε4 carriers predominantly show reduced performance in episodic memory, executive functioning, and, more marginally, perceptual speed, as compared to non-carriers. This has, however, produced findings that are difficult to interpret across studies because of variable methodology regarding the age groups involved, the cognitive measures employed, sample sizes, and study designs.

The precise role of *APOE* ε4 genotype on cognitive functioning therefore remains uncertain. A recent systematic review on the effect of APOE ε4 on cognition in the healthy population (O’Donoghue et al., 2018) suggested that it is challenging to disentangle cognitive deficits shown by APOE ε4 carriers in early AD pathology (‘Prodromal hypothesis’; Foster et al., 2013; Smith et al., 1998) from subtle cognitive deficits related to the APOE ε4 genotype (‘Phenotype hypothesis’; Fouquet et al., 2014; Greenwood et al., 2005; Parasuraman et al., 2002). While the former hypothesis predicts small to very small effect sizes on cognition in non-demented APOE ε4 carriers since any detectable effects would be due to cases of prodromal dementia (see Foster et al., 2013), the latter postulates that APOE ε4 carriers would show cognitive deficits that would be somehow independent of the development of AD due to interactions between APOE status and neuronal insult accumulated throughout the lifetime (see Greenwood et al., 2005; Payton et al., 2006). However, it is difficult to differentiate the relative importance of prodromal from phenotypic factors, and the evidence supporting the role of *APOE* genotype on cognitive abilities in the healthy population and the translational potential of this line of research remains still limited.

### 1.2 Episodic and Semantic Memory

Declarative or explicit memory refers to memories that can be consciously accessed and includes memory of specific lived events (episodic memory) and general knowledge of the world (semantic memory). While episodic memory entails re-experiencing and recollecting past events that are traceable in time and space (e.g., my 18^th^ birthday party in Montreal), semantic memory relates to conceptual knowledge abstracted over multiple experiences but detached from its context of acquisition (e.g., the definition of “birthday party” and knowledge of events that typically happen at birthday parties; Renoult et al., 2019).

When considering research on episodic and semantic memory in *APOE* ε4 carriers, existing studies have predominantly focused on episodic memory (see O’Donoghue et al., 2018; Small et al., 2004; Wisdom et al., 2011). This may be because episodic memory deficits are regarded as the early cognitive hallmark of Alzheimer’s Disease, where patients are commonly known to be impaired in the recollection of recent episodic events (McKhann et al., 2011). In a systematic review examining the role of the *APOE* ε4 genotype on episodic memory in AD patients, El Haj et al. (2016) indeed observed that most studies reported a significant relationship between *APOE* ε4 and episodic memory decline. The most recent meta-analysis available in the field (Wisdom et al., 2011) indicated that healthy ε4 carriers perform significantly worse on episodic memory and executive functioning tasks, in line with a previous meta-analysis (Small et al., 2004).

The distinction between episodic and semantic memory has also been questioned by studies that documented how these two forms of memory could be interdependent and overlapping in their neural correlates (see Greenberg & Verfaellie, 2010; Irish & Grilli, 2024; Tanguay et al., 2024). This distinction has also been revisited through evidence involving clinical populations (Buckley et al., 2014; Duval et al., 2012; Irish et al., 2010; Strikwerda-Brown et al., 2019). Semantic memory has been dissociated into personal and general semantics (Renoult et al., 2012; 2020 Grilli & Verfaellie, 2014, 2016; Strikwerda-Brown et al., 2019), with the former referring to knowledge of one’s personal past and the latter to wider culturally shared knowledge (e.g., vocabulary, maths, history, geography, uses of objects, knowledge of public events and famous people; Binder & Desai, 2011, Kumar, 2021; Reilly et al., 2025). Personal semantics has been operationalized in different ways across studies such as autobiographical facts (“I was born in 1982 in Alberta”), memory for repeated events (“I always celebrated my birthday at grandma’s when we lived in Canada”), and self-knowledge (“I am outgoing”). While these forms of personal semantics are traditionally included as part of semantic memory, recent studies have shown that the similarity between general and personal semantics (and with episodic memory) varied along with these different operationalizations Melega et al., 2024; Renoult et al., 2012, 2016; Tanguay et al., 2018; 2023; Grilli et al., 2018a; Grilli & Verfaellie, 2014, 2016; Marquine et al., 2016). Autobiographical memory is generally defined as including personal semantics and episodic memory (for a review, see Fan et al., 2024).

Despite these recent new insights, the role of semantic memory in healthy people at increased genetic risk of developing AD is yet to be clarified. Semantic memory was initially thought to be relatively spared at the earliest stages of the disease, as seen in famous case studies (see Gabrieli et al., 1988; O’Kane et al., 2004; Warrington & McCarthy, 1988) and less sensitive to aging (Nyberg et al., 2003), therefore consolidating the assumption that semantic memory may not be a sensitive marker for late-onset AD. A line of evidence has however challenged this view (Duff et al., 2020; Hoffman & Morcom, 2018; Verma & Howard, 2011), with cross-sectional studies involving people with Mild Cognitive Impairment (MCI) and AD which documented semantic memory impairments when using verbal fluency, naming and other similar tasks (Chasles et al., 2020; Joubert et al., 2010, 2021; Koenig et al., 2007; Storandt, 2008; Taler et al., 2016, 2020). Interestingly, in a study assessing autobiographical narratives in people with MCI and relative controls, Buckley and colleagues (2014) reported that personal semantic memory performance was related to beta-amyloid burden after adjusting for age and *APOE* ε4 genotype. In healthy APOE ε4 carriers, longitudinal studies including measures of semantic memory have however reported mixed results (Nilsson et al., 2006; Wilson et al., 2002), therefore the impact of the *APOE* ε4 genotype on semantic memory is still unclear.

### 1.3 Aims of the present review

To our knowledge, no previous systematic review has selectively investigated the impact of *APOE* ε4 on semantic memory in healthy adults. As evidence has suggested that semantic memory could be impaired in MCI and AD (e.g., Chasles et al., 2020; Joubert et al., 2010, 2020; Taler et al., 2016, 2020), but has largely been neglected in healthy people at increased genetic risk of developing AD, we aimed to review the available literature to scrutinize studies that reported and compared performance on semantic memory tasks in non-clinical adult populations with and without *APOE* ε4.

## 2. Methods

The initial search was carried out on 1^st^ March 2024 according to Preferred Reporting Items for Systematic reviews and Meta-Analyses (PRISMA) guidelines followed by an update search on 1^st^ September 2024, and another one carried out on 1^st^ March 2025. The search protocol and inclusion/exclusion criteria were pre-registered on the PROSPERO database (ID: CRD42024499684). Given the vast heterogeneity of the data and relatively small number of studies available (see section 3.3 Study Details), we adopted a narrative synthesis approach for this systematic review, as outlined by Popay et al. (2006).

### 2.1 Search strategy

The search strategy included the electronic databases: Academic Search Complete, AMED (The Allied and Complementary Medicine Database), CINAHL Complete (Cumulative Index of Nursing and Allied Health Literature), APA PsycArticles, APA PsycInfo, and MEDLINE Complete. The following search terms were used: “*APOE*” OR “*apolipoprotein*” AND “*memory*”)^1^.

As in previous reviews in the field (O’Donoghue et al., 2018), we only considered papers published from 1993, the year when *APOE* ε4 was first identified as a risk factor for AD (Corder et al., 1993). We also carried out a manual search by looking at reference lists of the articles included, systematic reviews, or meta-analyses relevant to the review topic.

### 2.2 Inclusion and exclusion criteria

The inclusion criteria were selected using the Population, Intervention, Comparison, Outcomes and Study (PICOS) framework (Methley et al., 2014; Pollock & Berge, 2018):

1. Population: healthy adults over the age of 18 without a diagnosis of neurodegenerative disease (including mild cognitive impairment), acquired brain injuries, psychiatric conditions, or reports of subjective memory complaints or decline;
2. Comparison: studies needed to report *APOE* genotype (i.e., ε2, ε3, ε4, or ε4 carriers vs non-carriers), and include a group comparison of heterozygous and/or homozygous *APOE* ε4 carriers versus non-carriers on semantic memory performance;
3. Outcome: Semantic memory performance assessed through standardized neuropsychological, cognitive, or experimental memory tasks;
4. Study: Empirical studies published in the English language.

In this process, we also referred to the following exclusion criteria:

1. Studies only including a paediatric population (under the age of 18);
2. Non-human animal studies;
3. Studies that did not report semantic memory performance at baseline (e.g., longitudinal study) and/or that did not mention semantic memory;
4. Reviews (including systematic reviews), meta-analyses, book chapters, and case reports;
5. Studies published in other languages than English;

### 2.3 Screening and Selection

Relevant articles were screened by title, abstract, and full-text after the removal of duplicates by the first reviewer (R.S.). A second reviewer (T.J.) screened 10% of the articles for the title and abstract and 20% of the articles for full-text. The second reviewer was randomly assigned a selection of articles to screen and was blind to the ratings of the first reviewer (R.S.). For both stages, the two reviewers discussed and resolved diverging views around inclusion or exclusion of papers.

### 2.4 Quality Rating

Quality assessment and critical appraisal of included articles was conducted using the Appraisal tool for Cross-Sectional Studies (AXIS – Downes et al., 2016). The AXIS tool includes 20 items with “Yes”, “No”, or “Not known” responses concerning the quality of reporting and of the study design, as well as potential sources of bias. The rating of risk of bias (“High”, “Medium” or “Low”) was based on reviewers’ judgment. To aid the quality rating process, a numerical rating was also computed: “Yes” answers received a point, and a “No” or “Not known” answer was scored as zero (excluding items 13 and 19, for which scores were reversed to “Yes/Not known ” = 0, “No” = 1).

As in the screening and selection process, the two reviewers completed this step and were blind to each other ratings. The second reviewer assessed the quality and risk of bias of approximately 50% of the included papers. Once the quality rating was completed, they discussed and resolved diverging views regarding the quality rating of the articles. The two raters agreed on almost all the items (154/160, 96.25%) and were able to resolve any disagreements.

## 3. Results

### 3.1 Study selection

Figure 1 shows the review process via the PRISMA 2020 flowchart diagram. The initial search from all the databases produced 7,881 articles. A total of 4,683 duplicates were removed, and a preliminary screening of 3,198 papers by title and abstract was completed. Forty-eight studies underwent full-text screening.

**Figure 1.**
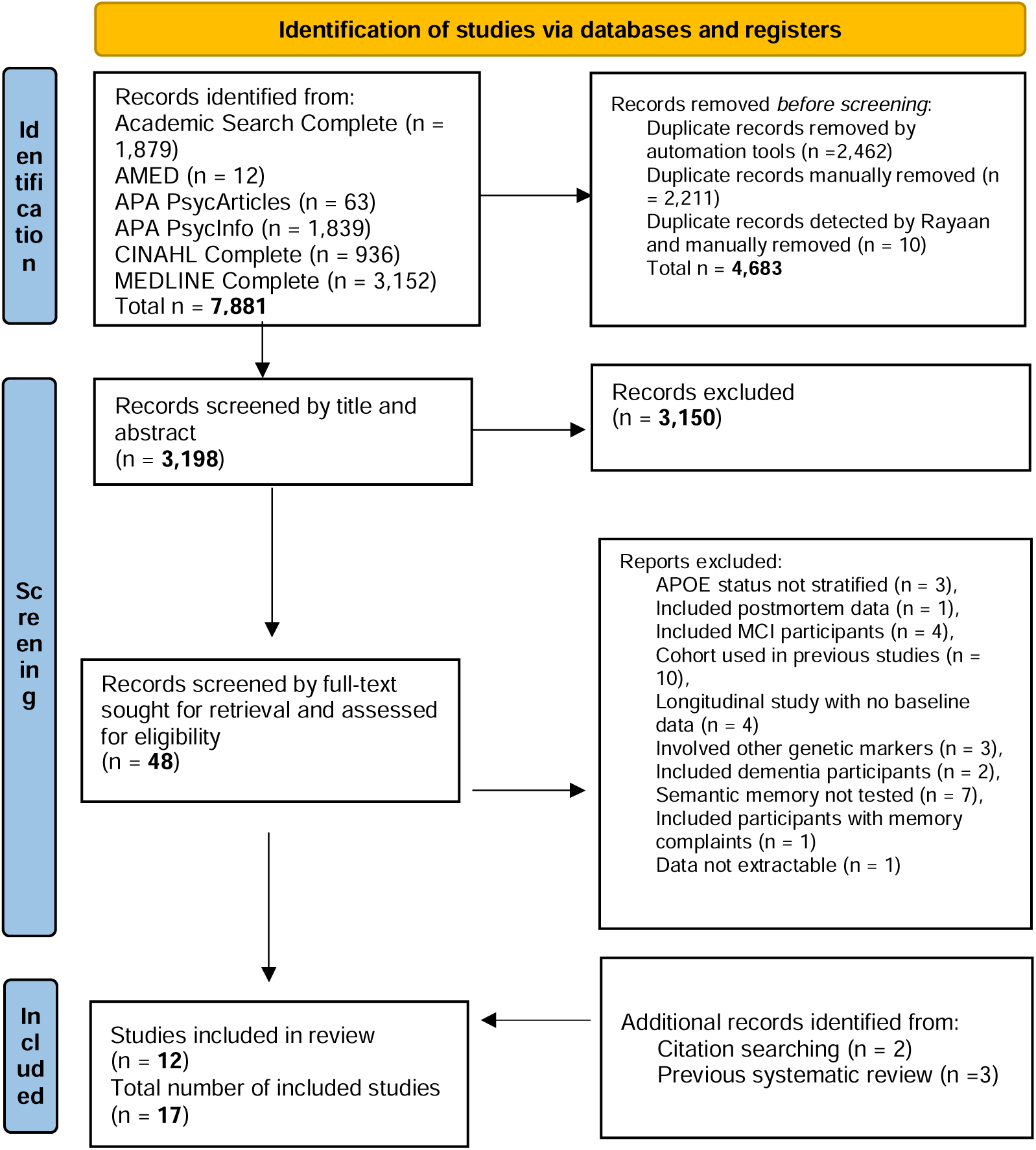
PRISMA flowchart outlining the article identification, screening and selection process.

We excluded 36 research articles during full-text screening (See Figure 1). This left 12 articles, all conventionally identified via databases. Two additional papers were identified via citation-searching of relevant papers, while three other papers were included in a previous systematic review on the effects of the APOE genotype on cognition (O’Donoghue et al., 2018). Seventeen papers were selected, with a total number of 8,491 participants tested.

### 3.2 Quality Assessment and Risk of Bias

Seven studies were rated as having “Medium” risk of bias, one paper was rated as “Medium to High” risk, and the remaining nine articles were considered to have a “Low” risk. Of the 17 articles, 10 did not justify the sample size, nor mentioned power analysis (Item 3).

### 3.3 Study Details

Detailed characteristics of each of the included studies are reported in *Table 1*. Apart from a single longitudinal study (Nilsson et al., 2006), all were cross-sectional studies including group comparisons between *APOE* ε4 carriers and non-carriers at a single time point. One paper (Seidenberg et al., 2009) also grouped the participants by family history for AD and *APOE* genotype to determine risk, while five papers stratified the participants by *APOE* genotype groups (i.e., *APOE* ε2/2, ε2/3, ε3/3, ε4/4, ε2/4, ε3/4; Helkala et al., 1995; Nilsson et al., 2006; Salo et al., 2001; Staehelin et al., 1999; Wikgren et al., 2012). The remaining 11 papers divided their participants between *APOE* ε4 carriers (+) and non-carriers (-).

**Table 1.** Tabulated results of the papers included in the systematic review.

| Author, Year | Study type | Sample size | Age groups | APOE groups | Semantic memory Task | Key finding/APOE ε4 effect | Risk of bias |
| --- | --- | --- | --- | --- | --- | --- | --- |
| Duchek et al., (2006) | Cross-sectional | n = 76 | Healthy Younger adults (18-24 years),<br>Young-old adults (65-78 years),<br>Old-old adults (80-93 years) | APOE ε4 (+)<br>APOE ε4 (-) | Information (WAIS-IV)<br>General Knowledge test (Einstein et al, 1995); Boston Naming Test (Kaplan et al., 1983); Animal Naming Test (Goodglass & Kaplan, 1983) | Higher performance on Animal Naming Test in Young-old<br>APOE ε4 (+) ($p = .013$, $d = 1.14$) | Medium |
| Eich et al., (2019) | Cross-sectional | n = 146 | Healthy Young and middle-aged adults (20-60 years) | APOE ε4 (+)<br>APOE ε4 (-) | Synonyms and Antonyms (Salthouse, 1993a,b); Picture Naming (Woodcock et al., 1989) | No significant group differences | Low |
| Ford et al., (2020) | Cross-sectional | n = 699 | Healthy older adults (60 - 85 years) | APOE ε4 (+)<br>APOE ε4 (-) | Categorization task (Stern & White, 2003) | Lower semantic clustering in APOE ε4 (+) ($p = .015$, $d = 0.22$) | Medium |
| Grilli et al., (2018b) | Cross-sectional | n = 40 | Healthy middle-aged and older adults (52-80 years) | APOE ε4 (+)<br>APOE ε4 (-) | Verbal Comprehension Index (WAIS-IV); Boston Naming Test (Kaplan et al., 1983)<br>Category Fluency Test<br>Autobiographical Memory Interview (Levine et al., 2002) | No significant group differences | Low |
| Grilli et al., (2021) | Cross-sectional | n = 45 | Healthy middle-aged and older adults (53 – 84 years) | APOE ε4 (+)<br>APOE ε4 (-) | Verbal Comprehension index (WAIS-IV)<br>Boston Naming Test (Kaplan et al., 1983)<br>Category Fluency test (COWAT, Benton, 1969)<br>Autobiographical fluency task (Addis & Tippett, 2004) | APOE ε4 (+) generated fewer exemplars on autobiographical fluency ($p = .02$, $\eta^2 = .13$), with lower personal semantic ($p = .02$, $d = .71$) and episodic memory fluency ($p = .02$, $d = .64$) | Medium |
| Helkala et al., (1995) | Cross-sectional | n = 916 | Healthy older adults (> 65) | APOE ε2/2,<br>ε2/3<br>APOE ε3/3<br>APOE ε4/4,<br>ε2/4, ε3/4 | Category Fluency | No significant group differences | Medium |
| Knoff et al., (2024) | Cross-sectional | n = 84 | Healthy middle-aged and older adults (60 – 80 years) | *APOE* ε4 (+)<br>*APOE* ε4 (-) | Verbal Comprehension index (WAIS-IV); Boston Naming Test (Kaplan et al., 1983); Category Fluency test | No significant group differences | Low |
| Laukka et al., (2013) | Cross-sectional | n = 2694 | Healthy older adults (60 - 90+ years) | *APOE* ε4 (+)<br>*APOE* ε4 (-) | SRB vocabulary test (Dureman, 1960)<br>General Knowledge task (Dahl et al., 2009) | No significant group differences | Low |
| Nilsson et al., (2006) | Longitudinal (Betula study) | n = 1733 | Middle-aged adults (35-50 years), Young-old adults (55-65 years), Old-old adults (70-85 years) | *APOE* ε3/3<br>*APOE* ε3/4<br>*APOE* ε4/4 | SRB vocabulary test (Dureman, 1960); Category Fluency | No significant group differences | Low |
| Payton et al., (2006) | Cross-sectional | n = 766 | Middle-aged and older adults (50 – 85 years) | *APOE* ε4 (+)<br>*APOE* ε4 (-) | Raven Mill Hill vocabulary scale parts A and B (Raven, 1965) | No significant group differences | Medium to High |
| Rosen et al., (2005) | Cross-sectional | n = 40 | Healthy middle-aged and older adults (50–79 years) | *APOE* ε4 (+)<br>*APOE* ε4 (-) | Extensive Category Fluency task (10 minutes); Category Fluency task (1-minute); Vocabulary (WAIS-IV) | *APOE* ε4 (+) generated fewer animal names ($p = .02$, $d = .68$), and fewer clusters of semantically related words ($p = .03$, $d = .63$) on extensive Category Fluency test and showed longer between-cluster retrieval times ($p = .03$, $d = .62$) | Medium |
| Salo et al., (2001) | Cross-sectional | n = 46 | Healthy older adults (> 85) | *APOE* ε2/2,2/3<br>*APOE* ε3/3<br>*APOE* ε4/4,<br>ε2/4, ε3/4 | Category Fluency test, Similarities Test WAIS-R | No significant group differences | Medium |
| Sapkota et al., (2016) | Cross-sectional | n = 282 | Healthy older adults (> 60 years) | *APOE* ε4 (+)<br>*APOE* ε4 (-) | Vocabulary task (Ekstrom et al., 1976) | No significant group differences | Low |
| Seidenberg et al., (2009) | Cross-sectional | n = 69 | Healthy older adults (65-85 years) | Control: No AD family history, *APOE* ε4 (-)<br>Group 1: AD Family history, *APOE* ε4 (-)<br>Group 2: AD family history, *APOE* ε4 (+) | Fame judgement task (Douville et al., 2005). | No significant group differences | Low |
| Stahelin et al., (1999) | Cross-sectional | n = 332 | Healthy older adults (> 65): Young-old (< 75 years)<br>Old -old (> 75 years) | *APOE* ε2/2, ε2/3, ε2/4<br>*APOE* ε3/3<br>*APOE* ε4/4, ε3/4 | Vocabulary (WAIS-R) | Higher performance in *APOE* ε3 group than *APOE* ε4 group ($p = .041$, $d = 0.29$) and trend for higher performance in *APOE* ε2 group compared to *APOE* ε4 group ($p = .062$, $d = 0.33$) | Medium |
| Tse et al., (2010) | Cross-sectional | n = 96 | Healthy Older adults (> 60 years) | *APOE* ε4 (+)<br>*APOE* ε4 (-) | Category Fluency<br>Information and Similarities (WAIS-IV) | No significant group differences | Medium |
| Wikgren et al., (2012) | Cross-sectional | n = 427 | Healthy middle-aged and older adults (41-85 years) | *APOE* ε3/3<br>*APOE* ε3/4<br>*APOE* ε4/4 | Revised version of the SRB vocabulary test (Dureman & Salde, 1971) | No significant group differences | Low |

The study sample sizes varied extensively, from samples of a few dozen participants (e.g., Grilli et al., 2018b, 2021; Rosen et al., 2005; Salo et al., 2001) to large cohorts of hundreds or even thousands of respondents (e.g., Ford et al., 2020; Helkala et al., 1995; Laukka et al., 2013; Nilsson et al., 2006; Payton et al., 2006).

Likewise, the age groups of the samples included in the studies varied too. All but one study included healthy older adults in their samples (Eich et al., 2019). Out of those 16 studies that included healthy older adults, three papers stratified the age of their participants by Young-Old, or Old-Old adults (e.g., < 75 years, and > 75 years respectively; Duchek et al., 2006; Nilsson et al., 2006; Stahaelin et al., 1999).

Eight studies also included middle-aged adults (i.e., between 40 to 60 years of age; Eich et al., 2019; Grilli et al., 2018b, 2021; Knoff et al., 2024; Nilsson et al., 2006; Payton et al., 2006; Rosen et al., 2005; Wikgren et al., 2012) and two studies also provided data from younger adults (i.e., between 18 to 35 years of age; Duchek et al., 2006; Eich et al., 2019).

The main source of heterogeneity among the selected studies derived from the type of test or task used to measure semantic memory. As outlined in *Table 2*, 11 studies adopted verbal fluency tasks (i.e., category fluency; Duchek et al., 2006; Ford et al., 2020; Grilli et al., 2018b, 2021; Helkala et al., 1995; Knoff et al., 2024; Nilsson et al., 2006; Rosen et al., 2005; Salo et al., 2001; Tse et al., 2010; Wikgren et al., 2012), five used naming tests (e.g., Boston Naming Test; Duchek et al., 2006; Eich et al., 2019; Grilli et al., 2018b, 2021; Knoff et al., 2024), and 15 used tests of language comprehension or general knowledge tests (e.g., verbal comprehension tests; Duchek et al., 2006; Eich et al., 2019; Grilli et al., 2018b, 2021; Knoff et al., 2024; Laukka et al., 2013; Nilsson et al., 2006; Payton et al., 2006; Rosen et al., 2005; Salo et al., 2001; Sapkota et al., 2016; Seidenberg et al., 2009; Stahaelin et al., 1999; Tse et al., 2010; Wikgren et al., 2012). Two studies assessed semantic memory by looking at autobiographical memory retrieval (Grilli et al., 2018b, 2021).

**Table 2.**
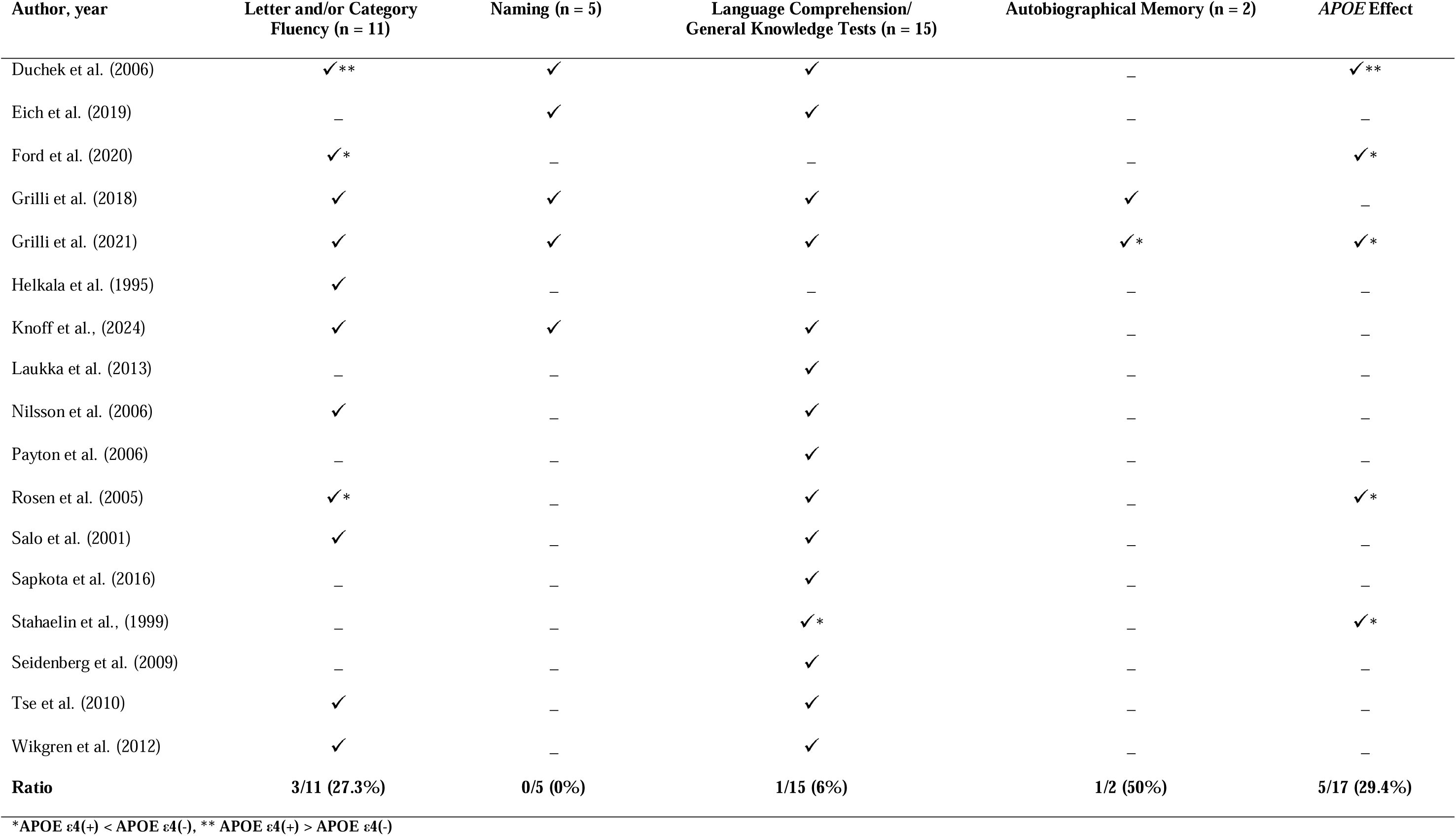
Findings of the selected papers tabulated by task used to measure Semantic Memory.

Given this heterogeneity in the methodology and tasks employed that may tap into different aspects of semantic memory as well as other cognitive abilities, we herein separately report the findings by the type of task used to measure semantic memory.

#### 3.3.1 Verbal Fluency

Verbal fluency tasks involve naming as many components of a particular semantic category (e.g., animals, fruits, vegetables), or as many words starting with a specific letter (e.g., F,A,S) in a specific time frame (usually one minute). The former task is typically referred to as Category or Semantic Fluency, and the latter as Letter Fluency. In these tasks, participants are typically warned against repeating the same word more than once, or in Letter Fluency, generating proper nouns, like names of people or places (e.g., cities, countries, regions). Tests of verbal fluency primarily assess the ability of accessing and retrieving words and their associations from an internal lexicon (Salthouse, 1991), as well as self-monitoring, and mental flexibility which are commonly referred to as Executive Functions (de Frias et al., 2005; Lezak et al., 2008). Given that Letter Fluency does not place explicit demand on semantic knowledge to be performed, only data from Category Fluency are included here.

Eleven studies included in this review considered category fluency tests as assessing semantic memory. Eight studies did not find any significant group differences using these tests (Grilli et al., 2018b, 2021; Helkala et al., 1995; Knoff et al., 2024; Nilsson et al., 2006; Salo et al., 2001; Tse et al., 2010; Wikgren et al., 2013). Two studies observed that *APOE* ε4 carriers performed significantly worse than non-carriers (Ford et al., 2020; Rosen et al., 2005). Finally, one study (Duchek et al., 2006) reported a reverse effect, with significantly higher performance among Young-Old *APOE* ε4 carriers (65 to 78 years of age) than non-carriers of the same age on a Category fluency task (*p* = .013, *d* = 1.14). The explanation of such reversed effects of *APOE* on cognition was not clear and could be a false positive finding (see Discussion section).

Along with the traditional one-minute Category fluency test, Rosen et al. (2005) also administered an extensive Category fluency task, where participants were asked to generate names from the animal category for 10 minutes and were also encouraged to generate names from subcategories (e.g., pets). Despite not finding any significant group differences in the one-minute Category fluency test, the authors reported that, in the 10 minutes version of the Category fluency task, *APOE* ε4 carriers generated fewer animal names (*p* = .02, *d* = .68), and fewer clusters of semantically related words (*p* = .03, *d* = .63), as compared to non-carriers. These participants also showed longer retrieval times when shifting from one semantic cluster to another, as compared to non-carriers (*p* = .03, *d* = .62).

More recently, Ford et al. (2020) assessed the ability of participants to generate groups of semantically similar items using the Categorization task (CAT; Stern & White, 2003), and to group words of similar meaning, as measured by the Semantic Clustering index (where a cluster corresponded to two or more words). The CAT task uses visual cues such as photographs and verbal information. While there were no significant differences in the Categorization task, the authors observed a lower semantic clustering performance in *APOE* ε4 carriers, compared to non-carriers (*p* = .015).

Considering the results of the category fluency tests together, it appears that *APOE* ε4 carriers’ performance on these tasks generally does not differ from the performance of non-carriers. This pattern of results does not seem to be influenced by the age groups of the participants involved, by the sample size included in the studies, or the rated risk of bias. However, those studies that employed a more complex variation of the verbal fluency tests reported a lower performance among *APOE* ε4 carriers (Ford et al., 2020; Rosen et al., 2005).

#### 3.3.2 Naming

Naming tests are designed to assess confrontational picture-naming and word retrieval and, more generally, expressive language. For instance, the commonly used Boston Naming Test (Kaplan et al., 1983) requires respondents to name a series of pictures of line-drawn objects and animals. If an object is not named spontaneously, participants are allowed to receive semantic cues (e.g., “something that contains water” for a glass). With naming abilities usually considered part of the language domain, the ability to recognise and name common objects largely draws upon the use of semantic knowledge and the lexicon.

In this review, five studies (Duchek et al., 2006; Eich et al., 2019; Grilli et al., 2018b, 2021; Knoff et al., 2024) included naming tests as a proxy measure of semantic memory, such as the Boston Naming Test (Kaplan et al., 1983), and the Picture Naming Test (Woodcock et al., 1989). As all five studies failed to detect any significant group difference (Duchek et al., 2006; Eich et al., 2019; Grilli et al., 2018b, 2021; Knoff et al., 2024), these findings thus suggest that the presence of *APOE* ε4 genotype does not generally seem to impact semantic memory when assessed through common language naming tasks. Crucially, some of these studies also reported the presence of ceiling effects in both carriers and non-carriers on the Boston Naming task (Duchek et al., 2006; Grilli et al., 2021; Knoff et al., 2024), as could be expected in samples of healthy older adults.

#### 3.3.3 Language Comprehension/General Knowledge Tests

Tests of language comprehension are also informative for semantic memory functioning. For instance, subtests of the Verbal Comprehension Index of the Weschler Adult Intelligence Scale (WAIS-IV; Weschler, 2008) are designed and standardised to assess understanding of language (e.g., Vocabulary), use of verbal reasoning (e.g., Similarities) and of verbal knowledge (e.g., Information), which all rely on semantic knowledge.

Fourteen studies included in this review employed a language comprehension task as a measure of semantic memory performance (Duchek et al., 2006; Eich et al., 2019; Grilli et al., 2018b, 2021; Knoff et al., 2024; Laukka et al., 2013; Nilsson et al., 2006; Payton et al., 2006; Rosen et al., 2005; Salo et al., 2001; Sapkota et al., 2016; Stahaelin et al., 1999; Tse et al., 2010; Wikgren et al., 2012). These tasks included the subtests of the Verbal Comprehension index of the WAIS-IV, the Synonym Reasoning Battery (SRB) Vocabulary Test (Dureman, 1960) and its revised version (Dureman & Salde, 1971), other Vocabulary tasks (see Ekstrom et al., 1976: Raven 1965), and Synonyms and Antonyms (Salthouse 1993a, 1993b).

Thirteen of these studies failed to detect a significant group difference in language comprehension tasks (Duchek et al., 2006; Eich et al., 2019; Grilli et al., 2018b, 2021; Knoff et al., 2024; Laukka et al., 2013; Nilsson et al., 2006; Payton et al., 2006; Rosen et al., 2005; Salo et al., 2001; Sapkota et al., 2016; Tse et al., 2010; Wikgren et al., 2013). Only Stahaelin et al. (1999) reported a significant effect of *APOE* ε4, whereby carriers performed significantly worse than non-carriers (*p* = .041, *d* = 0.29) on the Vocabulary test of the WAIS-Revised. Apart from this single study, the findings reported in the other studies predominantly suggest that, when semantic memory is measured through standard tests of language comprehension, *APOE* ε4 carriers and non-carriers do not seem to differ on these tasks. Nonetheless, in those studies including the Verbal Comprehension Index of the WAIS-IV (Grilli et al., 2018b, 2021; Knoff et al., 2024), mean composite scores suggested that participants tended to represent the high average range (110 to 119) or even in the superior range (120 to 129) of the general population, which indicates that these participants were highly educated for their age. This, therefore, may indicate a sampling bias and an inaccurate representation of the general population.

Tests of General Knowledge have also been frequently used as a measure of semantic memory (Bäckmann & Nilsson, 1996; Nyberg et al., 2003). These may include factual questions (“What is the capital of Paraguay?” or “What is the fastest animal in the world?”) or recognition questions, such as identifying the names or pictures of famous people (e.g., historical figures, politicians, actors, singers). In our systematic review, we included three papers using these types of tasks to assess semantic memory. Two studies (Duchek et al., 2006; Laukka et al., 2013) employed a General Knowledge Test (Dahl et al., 2009; Einstein et al., 1995), while Seidenberg et al., (2009) instead used a fame-judgement task, where carriers and non-carriers participants with and without an additional risk factor of a family history of AD were shown a series of names and were asked to rate them as “famous” or as “unfamiliar”. None of these studies observed any significant group differences between *APOE* ε4 carriers and non-carriers on task accuracy, or on reaction times. There were, however, indications of possible ceiling effects in Seidenberg et al., (2009), where participants’ performance in all groups exceeded 90% mean accuracy on the fame discrimination task, regardless of their genetic risk for developing AD (*APOE* genotype and family history).

#### 3.3.4 Autobiographical Memory

Semantic memory can also be measured via interview-based protocols that were developed to measure the retrieval of autobiographical memories. These include the Autobiographical Memory Interview (Kopelman et al., 1989) or the widely used Autobiographical Interview (Levine et al., 2002) and its more recent updated version (see Melega et al., 2024). These tasks are designed to assess and measure episodic and semantic memory retrieval, as they are both considered integrative parts of autobiographical memory.

In our systematic review, only two of the selected studies assessed the effect of *APOE* ε4 allele on semantic memory by considering autobiographical memory (Grilli et al., 2018b, 2021). In their first study, Grilli et al. (2018b) administered an adapted version of the Autobiographical Interview (Levine et al., 2002) to a group of *APOE* ε4 carriers and non-carriers. In this task, healthy older participants were asked to recall events from six different time periods, and detailed memory narratives for each life event were scored as internal (i.e., episodic) or external (including semantic details). While carriers produced autobiographical memories that were generally reduced in internal details as compared to non-carriers, Grilli et al. (2018b) did not observe any significant group difference in external details.

In a more recent study, Grilli et al. (2021) used an adapted version of the Autobiographical fluency tasks (Addis & Tippet, 2004; see also Dritschel et al., 1992) to assess episodic and personal semantic details. In this adapted task, participants were asked to generate exemplars of episodic (i.e., specific events) or personal semantic (e.g., names of personally relevant people) memories across three distinct life periods (childhood, early adulthood, recent life). Reportedly, *APOE* ε4 carriers generated fewer exemplars on this task than non-carriers, showing an overall lower fluency on personal semantic memory (*p* = .02, *d* = .71), as well as on episodic memory (*p* = .02, *d* = .64). Interestingly, *APOE* ε4 carriers did not show reduced performance in general semantic fluency tests, as measured by a standard neuropsychological test of category fluency (animals, fruits/vegetables). Based on these findings, the authors suggested that, along with reduced episodic memory, autobiographical memory deficits in *APOE* ε4 carriers could also extend to personal semantics, but not to general semantics.

Despite their very limited number, studies on autobiographical memory retrieval suggest that the presence of the *APOE* ε4 allele may not impact semantic memory when recollecting life events (Grilli et al., 2018b), or at least not all aspects of semantic memory, as it was observed in one study that *APOE* ε4 negatively impacted the generation of personal semantic memory, when it was assessed via the demands of an autobiographical fluency task (Grilli et al., 2021).

## 4. Discussion

As evidence has suggested that semantic memory could be impaired in MCI and AD (e.g., Chasles et al., 2020; Joubert et al., 2010, 2020; Taler et al., 2016, 2020), we aimed to systematically review the available literature that explored the role of *APOE* ε4 genotype on this memory domain in healthy adults at increased genetic risk of developing AD. Research in the field has abundantly reported episodic memory deficits associated with the *APOE* ε4 genotype (O’Donoghue et al., 2018; Small et al., 2004; Wisdom et al., 2011), while semantic memory has been more rarely investigated.

Overall, we found broad similarities in performance on semantic memory tasks between *APOE* ε4 and non-carriers, with some exceptions. The picture that, however, emerged from our systematic review is depicted by highly heterogeneous views on how semantic memory has been conceptualised and assessed over the past thirty years of research.

For instance, Nilsson et al. (2006) highlighted a theoretical ambiguity in how to classify verbal fluency tests. These authors critically stated that when relevant longitudinal studies in the field were commenced, verbal fluency tests were reliably regarded as tests of semantic memory (Backmann & Nilsson 1996; Nilsson et al., 1997), as they assessed the generation of words from an internal lexicon (Kausler, 1982, 1991), while they later started to be considered as part of a wider executive functioning assessment (de Frias et al., 2005; Salthouse et al., 2003). Similarly, even though naming tasks are often used as a measure of semantic memory, they are also employed as a measure of language production abilities. Even tasks assessing language or word comprehension that are considered more direct measures of semantic memory (Laukka et al., 2013; Nilsson et al., 2006), together with tasks assessing general knowledge of semantic facts (i.e., general semantics), still rely on other cognitive domains such as language and executive functioning. For consistency, we here briefly summarise the results of the effect of *APOE* ε4 genotype for each type of cognitive task used to assess semantic memory functioning.

When assessed with standard category fluency tasks, the studies here reviewed consistently reported similar semantic memory performance between *APOE* ε4 carriers and non-carriers, apart from one study from Duchek et al. (2006), where Young-Old *APOE* ε4 carriers outperformed non-carriers on an Animal Fluency task. To date, the paradoxical finding of improved performance in *APOE* ε4 carriers is not unusual in this research field (see Carrion-Baralt et al., 2009), as past studies also documented unaffected or even improved cognitive performance in *APOE* ε4 young adult carriers as compared to non-carriers of similar age (Acevedo et al., 2010; Bloss et al., 2010; Han & Bondi, 2008; Mondadori et al., 2007). Note however that the sample of participants showing a paradoxical effect of category fluency performance in Duchek et al. (2006) were 60+ and thus not typical of such reversed age effects. Moreover, there is still quite limited longitudinal evidence to support this hypothesis of varying *APOE* ε4 with age (see Ihle et al., 2012), and a recent meta-analysis failed to observe any significant differences between young carriers and non-carriers on several cognition domains (see Weissberger et al., 2018).

A significant genotype effect was detected in studies that employed a more complex version of category fluency tasks, where semantic memory was assessed over longer periods (i.e., 10 minutes, see Rosen et al., 2005). It could be argued that these complex fluency tasks are associated with heavier demands on executive functions (Eich et al., 2010; Rosen et al., 2005) that are known to be affected in ε4 carriers (O’Donoghue et al., 2018; Small et al., 2004; Wisdom et al., 2011), as observed in early studies investigating semantic memory in AD where patients showed difficulties on semantic tasks requiring self-initiation (e.g., category fluency, see Henry et al., 2004; Nebes, 1989). Moreover, studies assessing the ability of grouping words with similar meaning in category fluency tasks (i.e., semantic clustering) also observed that the *APOE* ε4 genotype was associated with reduced performance (Ford et al., 2020; Rosen et al., 2005), in line with studies that documented a decline in the usage of semantic clustering from MCI to a final diagnosis of AD (Malek-Ahmadi et al., 2011; McLaughlin et al., 2014).

When semantic memory was tested with naming tasks, there were no significant group differences between *APOE* ε4 carriers and non-carriers. Nevertheless, three studies also reported the presence of ceiling effects in the commonly used Boston Naming Task (Duchek et al., 2006; Grilli et al., 2021; Knoff et al., 2024), which could be expected in tasks that were initially designed for clinical populations. This therefore raises the question as to whether these tasks would be appropriate and sensitive enough to assess semantic memory in the healthy adult population, although performance on naming abilities was generally found to decline in late adulthood (see Verhaegen & Poncelet, 2012).

Taken together, when semantic memory is assessed via naming tasks, the evidence in support of *APOE* ε4 genotype effects remains limited and confined to one single study (Staehelin et al., 1999), that reported significantly lower performance in two groups of healthy older *APOE* ε4 carriers (Young-Old, Old-Old) as compared to non-carriers of the same age, while the rest of the papers reviewed did not observe significant group differences.

Only two studies assessed semantic memory retrieval with autobiographical memory tasks. The study from Grilli et al. (2018b) was the first and, so far, the only one adopting the autobiographical memory interview to compare *APOE* ε4 cognitively healthy middle-aged and older carriers and non-carriers. Nonetheless, in the autobiographical interview, external details include general semantics, personal semantics, but also metacognitive statements, comments and repetitions, and details about off-topic events, and are thus not a pure measure of semantic processing, though semantic details often represent an important portion of the interview transcripts, especially in older adults (Renoult et al., 2020). Further studies should clarify whether ε4 carriers and non-carriers differ in semantic details specifically.

The emerging finding of a specific impact on personal semantics, highlighted by Grilli et al. (2021), suggests that personal and general semantic fluency may entail different task demands, whereby personal semantics involved the retrieval of personally known names or spatiotemporal context (i.e., lifetime periods) which also share some episodic qualities (see Renoult et al., 2012) and is thought to be supported by medial temporal lobe regions (Grilli & Verfaellie, 2014, 2016; Conway, 2005; Greenberg et al., 2009; Sheldon & Moscovitch, 2012). Similar findings were also reported by Buckley et al. (2014), where personal semantic memory performance was related to neocortical beta-amyloid burden after adjusting for age and *APOE* status. Neuroimaging studies also documented changes in brain anatomy and connectivity in medial temporal lobe regions in healthy *APOE* ε4 carriers (Donix et al., 2010; Gallagher & Koh, 2011; Machulda et al., 2011; Mishra et al., 2018; for reviews see also Habib et al., 2017; Kucikova et al., 2021). Despite the restricted number of studies looking at this, the results of Grilli et al. (2021) suggest that the *APOE* ε4 genotype may not only affect episodic memory, but may be associated with broader autobiographical memory alterations including personal semantics.

Considering the overall results of this systematic review, most of the reviewed studies (70%) did not report significant group differences in semantic memory between *APOE* ε4 carriers and non-carriers. These findings are consistent with a previous systematic review on the effects of the *APOE* genotype on cognition that also considered semantic memory (O’Donoghue et al., 2018), although this review only included four studies evaluating semantic memory. As such, the evidence reviewed here predominantly suggests that *APOE* ε4 genotype is unlikely to influence semantic memory retrieval, at least when this is measured and assessed via standard neuropsychological tasks (i.e., verbal fluency, naming, and language comprehension tasks). Some group differences emerged when semantic memory was assessed via modified and more complex versions of verbal fluency tasks, or when measuring semantic clustering (Rosen et al., 20005; Ford et al., 2020), or when using autobiographical memory tasks allowing to differentiate personal and general semantics (Grilli et al., 2021).

This pattern of results indicates that the effect of *APOE* ε4 genotype on semantic memory could be revealed with a more precise assessment of semantic memory functioning. As observed in studies involving people with amnesia that used more complex semantic tasks (see Duff et al., 2020 for a review), semantic memory deficits could be similar to those of episodic memory as both memory domains rely on medial temporal regions. These tasks could include word associate tests, including identifying synonyms and common collocates, (i.e., words that often follow the target in a phrase or sentence, like “sudden” and “noise”), word senses tasks (i.e., name all the meanings that a related to a word, like “bank” as a financial institution or the bank of a river), and word feature tasks (i.e., name all of the features of a word or a concept, like “it barks”, “it can be a pet”, “it has different breeds”, “it has four legs” for the word “dog”) (Klooster & Duff, 2015), or extensive naming tasks (Hilverman & Duff, 2021), fairy tales or Bible stories (Rosenbaum et al., 2009; Verfaellie et al., 2014) or even generating hypothetical meaning for novel word compounds (e.g., cactus carpet, see Keane et al., 2020). Moreover, longitudinal evidence also suggested that semantic memory performance may decline over time in ε4 carriers, when assessed through a composite score combining verbal fluency with naming, reading and vocabulary abilities (see Wilson et al., 2002). Nonetheless, as mentioned above, the impact of other factors such as increased demands on executive functions has to be considered in these more complex tasks. In the case of autobiographical memory tasks, such as autobiographical fluency (Addis & Tippet, 2004; Dritschel et al., 1992; Grilli et al., 2021), a contribution of episodic memory processes is also likely (Greenberg et al., 2009; Ryan et al., 2008; Sheldon & Moscovitch, 2012).

It is worth noting that every study included in this systematic review made use of verbal tasks. To better determine the impact of *APOE* ε4 on semantic memory, future research should also include non-verbal tasks in test batteries (e.g., semantic associations tasks using pictures and sounds; see Bozeat et al., 2000, 2003). There is also a clear need for more studies adopting measures of autobiographical memory. Such tasks and interview protocols could arguably represent a more ecologically valid assessment of episodic and semantic memory function, which are notionally linked to brain areas that are vulnerable to the early stages of AD pathology (i.e., medial temporal lobes, see Martinelli et al., 2013), as already stated for more complex general semantic tasks (Duff et al., 2020). Therefore, a more precise assessment of semantic memory is needed to better understand whether this cognitive domain is affected by *APOE* ε4 genotype, and studies focusing on autobiographical memory tasks could help cast light on this matter.

### 4.1 Other sources of heterogeneity and limitations

The age of the participants included in the selected papers could have also been a source of heterogeneity and bias in our findings, as most *APOE* ε4 effects on cognition are observed in older adults. Out of the 17 papers here reviewed, 16 included healthy older adults (94%), while 8 studies also included middle-aged adults (47%) and two even included younger adults (11.8%). Crucially, all the significant findings relating *APOE* ε4 to semantic memory were observed in studies involving cohorts of older adults. However, as almost all studies included older adults and only few considered participants in early adulthood, the mediating influence of age on *APOE* ε4 effects on semantic memory is still unclear.

The studies included in this review differed in terms of *APOE* genotype types used to allocate participants into groups, with thirteen studies considering overall group differences between *APOE* ε4 carriers and non-carriers (Duchek et al., 2006 Eich et al., 2019; Ford et al., 2020; ; Grilli et al., 2018b, 2021; Knoff et al., 2024; Laukka et al., 2013; Payton et al., 2006; Rosen et al., 2005; Sapkota et al., 2016; Seidenberg et al., 2009; Tse et al., 2010), while five stratified respondents for each genotype group (Helkala et al., 1995; Nilsson et al., 2006; Staehelin et al., 1999; Salo et al., 2001; Wikgren et al., 2012). By looking at the date of publication of these latter studies, it appears that they were mainly published in the early years of *APOE* genotype research (later 90s/early 2000s) when the ε4 genotype was still being investigated as a potential genetic risk factor for AD. Instead, later research in the field then started to compare samples with homozygote and heterozygote ε4 carriers to groups of participants who were simply considered non-carriers, likely because the evidence around the effect *APOE* ε4 on cognition has become more consolidated. Nonetheless, recruiting an adequate number of *APOE* ε4 homozygote carriers can also be quite challenging, as these participants are quite rare in the general population (see Caselli & Reiman, 2013), so recruitment would therefore require very large samples of participants, usually from already genotyped cohorts.

### 4.2 Conclusions

Considering recent research advances that have revisited the role of semantic memory in AD, we systematically reviewed studies comparing healthy adult *APOE* ε4 carriers and non-carriers on semantic memory tasks. Our findings indicate a pervasive heterogeneity and a lack of consensus on the conceptualisation and therefore the assessment of semantic memory. When tested via classic neuropsychological tests that mainly assess general semantic memory, the performance *APOE* ε4 carriers did not generally differ from non-carriers. When semantic memory was assessed via modified versions of verbal fluency tasks or considering semantic clustering, carriers were found to be impaired. Similarly, in one study considering retrieval fluency of autobiographical memories, carriers showed a deficit in the generation of personal semantic information, compared to non-carriers (Grilli et al., 2021).

We conclude that the impact of *APOE* ε4 on semantic memory may be restricted to more demanding tasks, which could constitute a better match to episodic memory tasks for which effects are typically observed (Small et al., 2004; Wisdom et al., 2011), though a mediating role of executive functions should also be considered (O’Donoghue et al., 2018; Wisdom et al., 2011). Future studies on autobiographical memory retrieval in *APOE* ε4 carriers could provide a more precise and ecologically valid assessment of semantic memory, especially when disentangling between personal and general forms of semantic memory.

## Authors’ contributions

This work was ideated by RS, JB and LR. Literature search and evaluation was completed by the first rater (RS) with the help of a second rater (TJ). The first draft was written by RS and all authors critically read and revised the work, and eventually approved the final manuscript.

## Declarations

None.

## Data availability

No data are available for this study other than the ones reported in this manuscript.

## Footnotes

1 We chose a broad search term (“memory”) to maximise sensitivity, as initial searches revealed that relevant studies did not always use the expression “semantic memory”.

